# Interpretable surface-based detection of focal cortical dysplasias: a MELD study

**DOI:** 10.1101/2021.12.13.21267721

**Authors:** Hannah Spitzer, Mathilde Ripart, Kirstie Whitaker, Antonio Napolitano, Luca De Palma, Alessandro De Benedictis, Stephen Foldes, Zachary Humphreys, Kai Zhang, Wenhan Hu, Jiajie Mo, Marcus Likeman, Shirin Davies, Christopher Guttler, Matteo Lenge, Nathan T. Cohen, Yingying Tang, Shan Wang, Aswin Chari, Martin Tisdall, Nuria Bargallo, Estefanía Conde-Blanco, Jose Carlos Pariente, Saül Pascual-Diaz, Ignacio Delgado-Martínez, Carmen Pérez-Enríquez, Ilaria Lagorio, Eugenio Abela, Nandini Mullatti, Jonathan O’Muircheartaigh, Katy Vecchiato, Yawu Liu, Maria Caligiuri, Ben Sinclair, Lucy Vivash, Anna Willard, Jothy Kandasamy, Ailsa McLellan, Drahoslav Sokol, Mira Semmelroch, Ane Kloster, Giske Opheim, Letícia Ribeiro, Clarissa Yasuda, Camilla Rossi-Espagnet, Kai Zhang, Khalid Hamandi, Anna Tietze, Carmen Barba, Renzo Guerrini, William Davis Gaillard, Xiaozhen You, Irene Wang, Sofía González-Ortiz, Mariasavina Severino, Pasquale Striano, Domenico Tortora, Reetta Kalviainen, Antonio Gambardella, Angelo Labate, Patricia Desmond, Elaine Lui, Terence O’Brien, Jay Shetty, Graeme Jackson, John Duncan, Gavin Winston, Lars Pinborg, Fernando Cendes, Fabian J. Theis, Russell T. Shinohara, J Helen Cross, Torsten Baldeweg, Sophie Adler, Konrad Wagstyl

**Affiliations:** Institute of Computational Biology, Helmholtz Center Munich, Munich, Germany; UCL Great Ormond Street Institute for Child Health, 30 Guilford St, Holborn, London WC1N 1EH; The Alan Turing Institute, 96 Euston Rd, Somers Town, London NW1 2DB; The Bambino Gesù Children’s Hospital, Medical Physics Department, Rome; The Bambino Gesù Children’s Hospital, Rome; Barrow Neurological Institute at Phoenix Children’s Hospital, 1919 E Thomas Rd, Phoenix, AZ 85016; Department of Neurosurgery, Beijing Tiantan Hospital, Capital Medical University, Beijing; Bristol Royal Hospital for Children, Upper Maudlin St, Bristol, UK; Cardiff University Brain Research Imaging Centre, School of Psychology, Cardiff; Cardiff and Vale University Health Board, The Welsh Epilepsy Unit, University Hospital of Wales, Cardiff, UK; Charité University Hospital, Berlin; Neuroscience Department, Children’s Hospital Meyer-University of Florence, viale Pieraccini 24, 50139, Florence, Italy; Center for Neuroscience Research, Children’s National Hospital, The George Washington University School of Medicine, Washington, DC, USA; Department of Neurology, West China Hospital of Sichuan University, Chengdu, China; Epilepsy Center, Cleveland Clinic, Cleveland, OH, USA; Department of Neurology, Epilepsy Center, Second Affiliated Hospital, School of Medicine, Zhejiang University, Hangzhou, China; Great Ormond Street Hospital NHS Foundation Trust, UK; Magnetic Resonance Imaging, Core Facility, IDIBAPS, C/ Rossello 149, Barcelona 08036, Spain; Department of Neurosurgery, Hospital del Mar, Barcelona; Department of Neurology, Hospital del Mar, Barcelona; IRCCS Istituto Giannina Gaslini, Via Gaslini 5, 16147, Genova, Italy; Center for Neuropsychiatry and Intellectual Disability, Psychiatrische Dienste Aargau AG, Königsfelderstrasse 1, 5120 Windisch, Switzerland; Institute of Psychiatry, Psychology and Neuroscience, 16 De Crespigny Park, Camberwell, London SE5 8AB; Department of Forensic and Neurodevelopmental Sciences, MRC Centre for Neurodevelopmental Disorders, Institute of Psychiatry, Psychology and Neuroscience, King’s College London; Department of Perinatal Imaging and Health, St. Thomas’ Hospital, King’s College London; Department of Forensic and Neurodevelopmental Sciences, Institute of Psychiatry, Psychology and Neuroscience, King’s College London; Department of Neurology, University of Eastern Finland, Kuopio, Finland; Neuroscience Research Center, Department of Medical and Surgical Sciences, Magna Graecia University of Catanzaro, Viale Europa, Catanzaro, Italy, 88100; Department of Neuroscience, Central Clinical School, Monash University, Level 6 The Alfred Centre, 99 Commercial Road, Melbourne, Victoria, 3004; Central Clinical School, Alfred Centre, 99 Commercial Rd, Melbourne VIC 3004; Department of Neurology, Alfred Health, Commercial Road, Melbourne, Australia; Royal Hospital for Children and Young People, Little France Crescent, Edinburgh EH16 4TJ; The Florey Institute of Neuroscience and Mental Health, Austin Campus, 245 Burgundy St, Heidelberg, VICTORIA 3071, Australia; Neurobiology Research Unit, 6-8 Inge Lehmanns Vej, Rigshospitalet, building 8057, DK-2100 Copenhagen, Denmark; Department of Neurology, University of Campinas - UNICAMP, Rua Vital Brasil, 251, Cidade Universitária, Campinas, SP, Brazil 13083-888; Brazilian Institute of Neuroscience and Neurotechnology (BRAINN), Campinas, SP, Brazil; Neuroradiology Unit, IRCCS Bambino Gesù Children’s Hospital, Rome; Department of Neurosurgery, Beijing Tiantan Hospital, Capital Medical University, Beijing, China; Cardiff University Brain Research Imaging Centre, School of Psychology, Cardiff, UK; The Welsh Epilepsy Unit, University Hospital of Wales, Cardiff; Center for Neuroscience, Children’s National Hospital, Washington DC USA; Department of Radiology, Hospital del Mar, Barcelona; Kuopio Epilepsy Center, Neurocenter, Kuopio University Hospital, Kuopio, Finland; Institute of Neurology, Department of Medical and Surgical Sciences, Magna Graecia University, Viale Europa, Catanzaro 88100, Italy; Department of Radiology, Royal Melbourne Hospital, University of Melbourne, Parkville, Victoria, Australia, 3050; Department of Radiology, Royal Melbourne Hospital, 300 Grattan St, Parkville, Victoria, Australia, 3050; Monash University, 99 Commercial Road, Melbourne, Victoria, 3004, Australia; Department of Medicine (The Royal Melbourne Hospital), Royal Parade, Parkville, Victoria, 3052; Department of Neurology, Austin Health, Heidelberg, Australia; UCL Queen Square Institute of Neurology, Queen Square, London, WC1N 3BG; Department of Medicine, Division of Neurology, Queen’s University, Kingston, Canada; Department of Neurology, University of Campinas, Rua Vital Brasil, 251, Cidade Universitária, Campinas, SP, Brazil 13083-888; Department of Mathematics, Technical University of Munich, Germany; Penn Statistics in Imaging and Visualization Center, Department of Biostatistics, Epidemiology, and Informatics, Perelman School of Medicine, University of Pennsylvania, Philadelphia, PA 19104 USA; Wellcome Centre for Human Neuroimaging, 12 Queen Square, Holborn, London WC1N 3AR

**Author notes:** **Corresponding author:** Konrad Wagstyl, Wellcome Centre for Human Neuroimaging, 12 Queen Square, London WC1N 3AR.

**Keywords:** Focal cortical dysplasia, epilepsy, structural MRI, machine-learning

## Abstract

**Introduction:** One outstanding challenge for machine learning in diagnostic biomedical imaging is algorithm interpretability. A key application is the identification of subtle epileptogenic focal cortical dysplasias (FCDs) from structural MRI. FCDs are difficult to visualise on structural MRI but are often amenable to surgical resection. We aimed to develop an open-source, interpretable, surface-based machine-learning algorithm to automatically identify FCDs on heterogeneous structural MRI data from epilepsy surgery centres worldwide.

**Methods:** The Multi-centre Epilepsy Lesion Detection (MELD) Project collated and harmonised a retrospective MRI cohort of 1015 participants, 618 patients with focal FCD-related epilepsy and 397 controls, from 22 epilepsy centres worldwide. We created a neural network for FCD detection based on 33 surface-based features. The network was trained and cross-validated on 50% of the total cohort and tested on the remaining 50% as well as on 2 independent test sites. Multidimensional feature analysis and integrated gradient saliencies were used to interrogate network performance.

**Results:** Our pipeline outputs individual patient reports, which identify the location of predicted lesions, alongside their imaging features and relative saliency to the classifier. Overall, after including a border-zone around lesions, the developed MELD FCD surface-based algorithm had a sensitivity of 67% and a specificity of 54% on the withheld test cohort, and a sensitivity of 85% on a restricted subcohort of seizure free patients with FCD type IIB who had T1 and FLAIR MRI data.

**Conclusions:** This multicentre, multinational study with open access protocols and code has developed a robust and interpretable machine-learning algorithm for automated detection of focal cortical dysplasias, giving physicians greater confidence in the identification of subtle MRI lesions.

**Highlights:**

- This large, multi-centre, multi-scanner neuroimaging cohort captures the heterogeneity of histopathological subtypes and imaging features of patients with FCD.
- We developed a robust and interpretable surface-based algorithm which detects FCDs with a sensitivity of 67% and a specificity of 54%.
- The algorithm generates individual patient reports that “open the AI black-box” highlighting predicted lesion locations, alongside the imaging features and their relative saliency to the classifier.

## 1. Introduction

The application of machine learning algorithms for diagnostics in biomedical imaging forms a spectrum from automating high-throughput imaging analysis to assisting diagnosis in rarer, clinically-challenging pathologies. One barrier to clinical translation is the limited interpretability of these algorithms, leading to a common perception of them as impenetrable “black-boxes”. Identifying focal epileptogenic abnormalities on MRI is an outstanding clinical challenge in patients undergoing presurgical evaluation for drug-resistant focal epilepsy (DRFE). In DRFE, 16-43% of individuals are “MRI-negative”, i.e. no relevant abnormality is visually identified on their MRI scans (Bien et al., 2009; Colombo et al., 2012; McGonigal et al., 2007). A leading cause of DRFE and the most common histopathology in operated “MRI-negative” cohorts is a malformation of cortical development, called focal cortical dysplasia (FCD) (Irene Wang et al., 2013). As post-surgical seizure freedom is affected by whether the FCD can be identified on preoperative structural MRI (Bien et al., 2009; Téllez-Zenteno et al., 2010), there has been considerable effort placed in improving the detection of these lesions. However, machine learning approaches provide little insight into factors determining classification. In clinically ambiguous images, where the need for algorithms is greatest, such insight would enable physicians to determine whether features identified by the classifiers are likely to be lesional in origin.

Radiologically FCDs are characterised by alterations in cortical thickness, blurring at the grey-white matter boundary, folding abnormalities, and T2 or FLAIR signal intensity changes (Colombo et al., 2012). Approaches to improving the detection of FCDs have involved improved scanner protocols (Bernasconi et al., 2019) and field strengths (Bartolini et al., 2019; Wang et al., 2020) as well as automated volumetric (David et al., 2021; Gill et al., 2021, 2018; House et al., 2021; Huppertz et al., 2005) and surface-based (Adler et al., 2017; Ahmed et al., 2015; Hong et al., 2014; Jin et al., 2018) post-processing methods.

Despite extensive retrospective work to improve FCD detection, few automated methods have been used prospectively in the presurgical evaluation of patients with epilepsy. Alongside lack of interpretability, there are many additional reasons for this. First, some of the frameworks have been developed at single epilepsy centres resulting in small sample sizes and homogeneous datasets, where all patients have been scanned on the same MRI scanner with the same protocol, which reduces the likelihood of robustness of the results and the ability of the method to generalise. Second, many of the frameworks are not openly available and therefore difficult to reproduce. Third, much of the research has been limited to “MRI-positive” patients or limited to histopathologically confirmed FCD type IIA or IIB patients and thus have unknown performance on many of the complex, challenging patients who present to epilepsy surgery centres. Although, there has been some important research replicating previous methods (Jin et al., 2018; Wagstyl et al., 2020; Wang et al., 2015), there is a need to develop and validate an automated FCD detection tool on a large multi-centre cohort and make it available to the epilepsy community.

Here, as part of the MELD Project (Wagstyl et al., 2021), we aimed to collate a cohort of patients from multiple epilepsy surgery centres, across multiple MRI scanners including both 1.5T and 3T field strengths; create protocols for de-centralised MRI post-processing; and develop an open-access, robust and interpretable surface-based classifier to detect FCD.

## 2. Methods

### 2.1 MELD project consortium

The MELD project (https://meldproject.github.io/) involves 22 research centres across 5 continents. Each centre received approval from their local institutional review board (IRB) or ethics committee (EC). IRB/EC waived the need for individual patient consent as this was a retrospective study using fully anonymised routinely available data only.

### 2.2 Participants

Patients were included if they were over age 3, had a 3D preoperative T1-weighted MRI brain scan (1.5T or 3T) and a radiological diagnosis of FCD or were MRI-negative with histopathological confirmation of FCD. Participants were excluded if they had previous surgery, large structural abnormalities in addition to the FCD or T1 scans with gadolinium-enhancement. Controls were included if they were over age 3, did not have epilepsy or another neurological condition and had a T1-weighted MRI brain scan (1.5T or 3T). Patients scanned for headache could be included as controls if they had no other neurological conditions and the MRI was normal. The patients and controls included were a retrospective convenience sample. Centres, patients and controls were given pseudo-anonymised ID codes.

### 2.3 Methods overview

Figure 1 is an overview of the MELD FCD processing pipeline, which is explained in more detail in the sections below.

**Figure 1.**
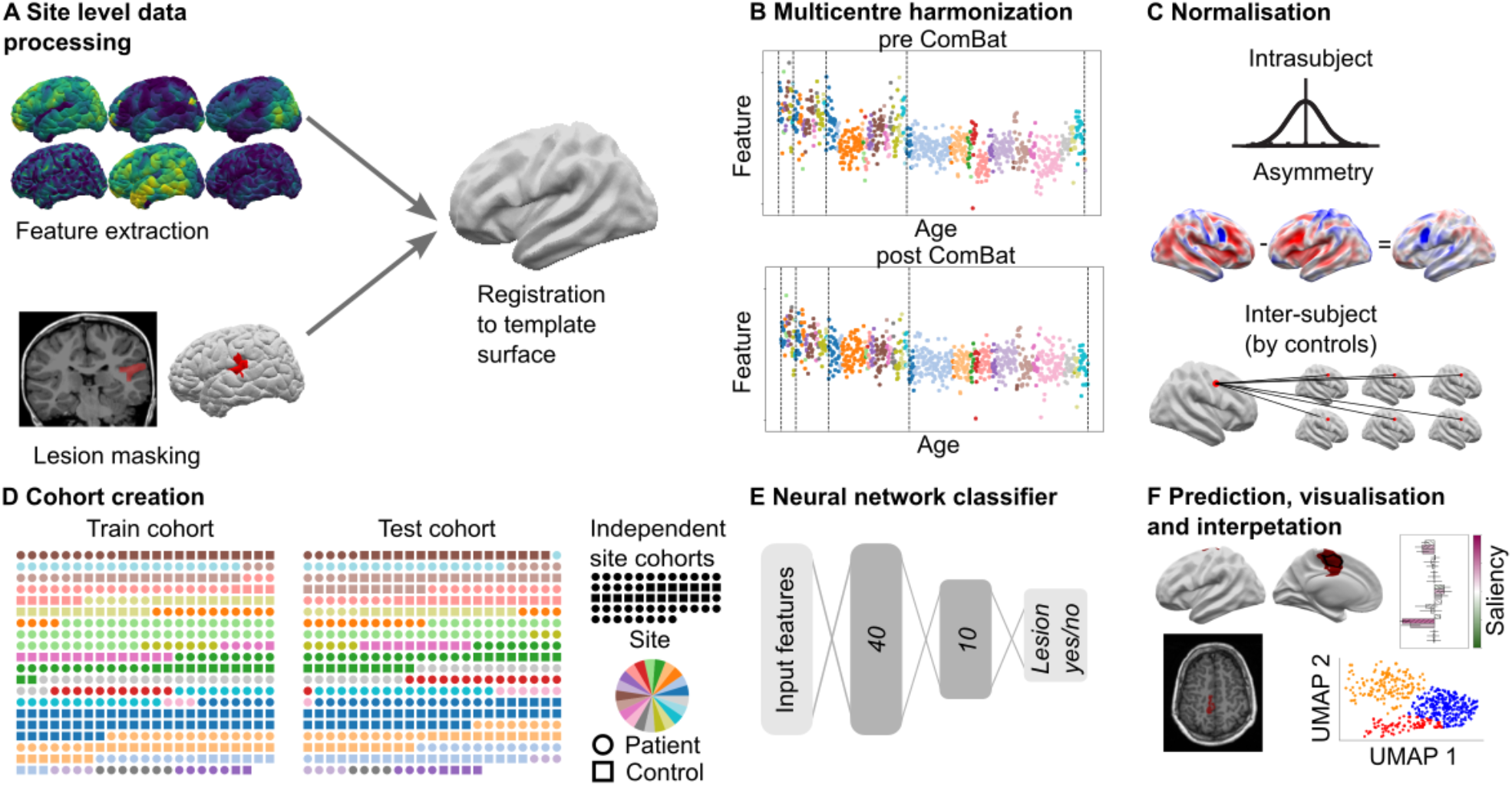
MELD processing pipeline. A) Local sites extract surface-based morphological features from structural T1 and FLAIR MRI, along with manually delineated lesion masks. These were coregistered to symmetric template surface and anonymised data matrices are shared with MELD team. B) Central preprocessing: MELD team carried out outlier detection and data harmonisation to minimise inter-scanner feature differences. C) Morphological features underwent intra-subject, interhemispheric and inter-subject normalisation. D) The full cohort was randomly subdivided 50:50 into training/validation cohorts and withheld test cohort. To avoid overfitting, all optimisation experiments were carried out on the train/val cohort prior to final testing on the test cohort and new site cohorts. E) Neural network classifier was trained to identify lesional vertices from MRI features. Vertex-wise predictions were collected into connected clusters. F) Classifier predictions mapped to cortical surfaces, lesional features and their relative saliency were plotted, lesional features across the cohort were analysed.

### 2.4 Site-level data collection and post-processing

Each site followed the protocols for site-level data collection and post-processing that are available at https://www.protocols.io/researchers/meld-project.

#### 2.4.1. Participant demographics

The following data were collected for all patients: age at preoperative scan, sex, age of epilepsy onset, duration of epilepsy (time from age of epilepsy onset to age at preoperative scan), ever reported MRI-negative and histopathological diagnosis (ILAE three-tiered classification system),(Blümcke et al., 2011) seizure-freedom (Engel class I or other) and follow-up time in operated patients.

Deidentified participant demographic information and lesion masks were shared with the MELD project coordinators for multi-centre analysis. No identifiable patient information was shared.

#### 2.4.2. MRI data collection and cortical surface reconstruction

3D T1-weighted and FLAIR (where available) MRI scans were collected at the 22 participating centres for all participants and cortical surfaces were reconstructed using *FreeSurfer*.*(Fischl, 2012)* Detailed protocols for structural MRI post-processing were adapted from openly available ENIGMA-epilepsy protocols.(Whelan et al., 2018)

#### 2.4.3. FCD lesion masking

FCD lesions were delineated on the T1-weighted MRI scans at each site according to our lesion masking protocol.(MELD Project, n.d.) For patients with a radiological diagnosis of FCD, a volumetric lesion mask was created using the preoperative T1 scan and 3D FLAIR (where available). For MRI-negative patients but with histopathological confirmation of FCD, the post-operative scan was used to create the volumetric lesion mask. In both cases, masks were performed by a neuroradiologist, neurologist, or experienced epilepsy researcher at each site. Volumetric lesion masks were mapped to cortical reconstructions and small defects were filled in using five iterations of a dilation-erosion algorithm. Patients’ lesions were registered to *fsaverage_sym*.

#### 2.4.4. Morphological/intensity features

The following measures were calculated in native space per vertex across the cortical surface in all participants: (a) cortical thickness, (b) contrast in T1 intensity above and below the grey-white matter boundary (grey-white contrast) (c) mean curvature, (d) sulcal depth, and (e) intrinsic curvature. In participants with FLAIR data, FLAIR signal intensity was sampled at 25%, 50%, and 75% of the cortical thickness (GM FLAIR 25%, 50%, 75%), as well as at the gray-white matter boundary and 0.5 and 1 mm subcortically (WM FLAIR 0.5mm, 1mm). The following features were smoothed with a 10mm Gaussian kernel: cortical thickness, intensity at the gray-white matter contrast, and FLAIR intensities at all cortical and subcortical depths, to increase the stability of per-vertex measures. Intrinsic curvature was smoothed with a 20mm Gaussian kernel to provide a measure of folding pattern abnormalities that is stable across adjacent gyri and sulci. All features were registered to bilaterally symmetrical template space, *fsaverage_sym*. Only anonymised participant demographic details and data matrices of anonymised features and lesion masks were shared with the central analysis site (UCL).

### 2.5 Centralised quality control and post-processing

#### 2.5.1. Quality control and data harmonisation of surface-based data

Automated quality control was performed on the surface-based features to identify subjects with extreme structural and intensity values across multiple features and cortical areas, likely caused by imaging artifacts such as signal biases or *FreeSurfer* segmentation errors. A feature was considered an outlier if, in more than 10 non-lesional regions (from the Desikan-Killiany atlas), it was greater or less than 2.7 times the standard deviation from the mean of all participants values. Participants were considered outliers if they had multiple extreme features, two if features from T1-weighted scans only and three if FLAIR MRI scans available. Participants identified as outliers were excluded from all subsequent analyses.

Structural and intensity features were harmonised across sites and scanners strength using ComBat(Fortin et al., 2018), controlling for age, sex and disease status (Figure 2). Independent test sites were harmonised to the main cohort (Figure 2). The harmonised dataset features are henceforth referred to as “ComBat” features.

**Figure 2.**
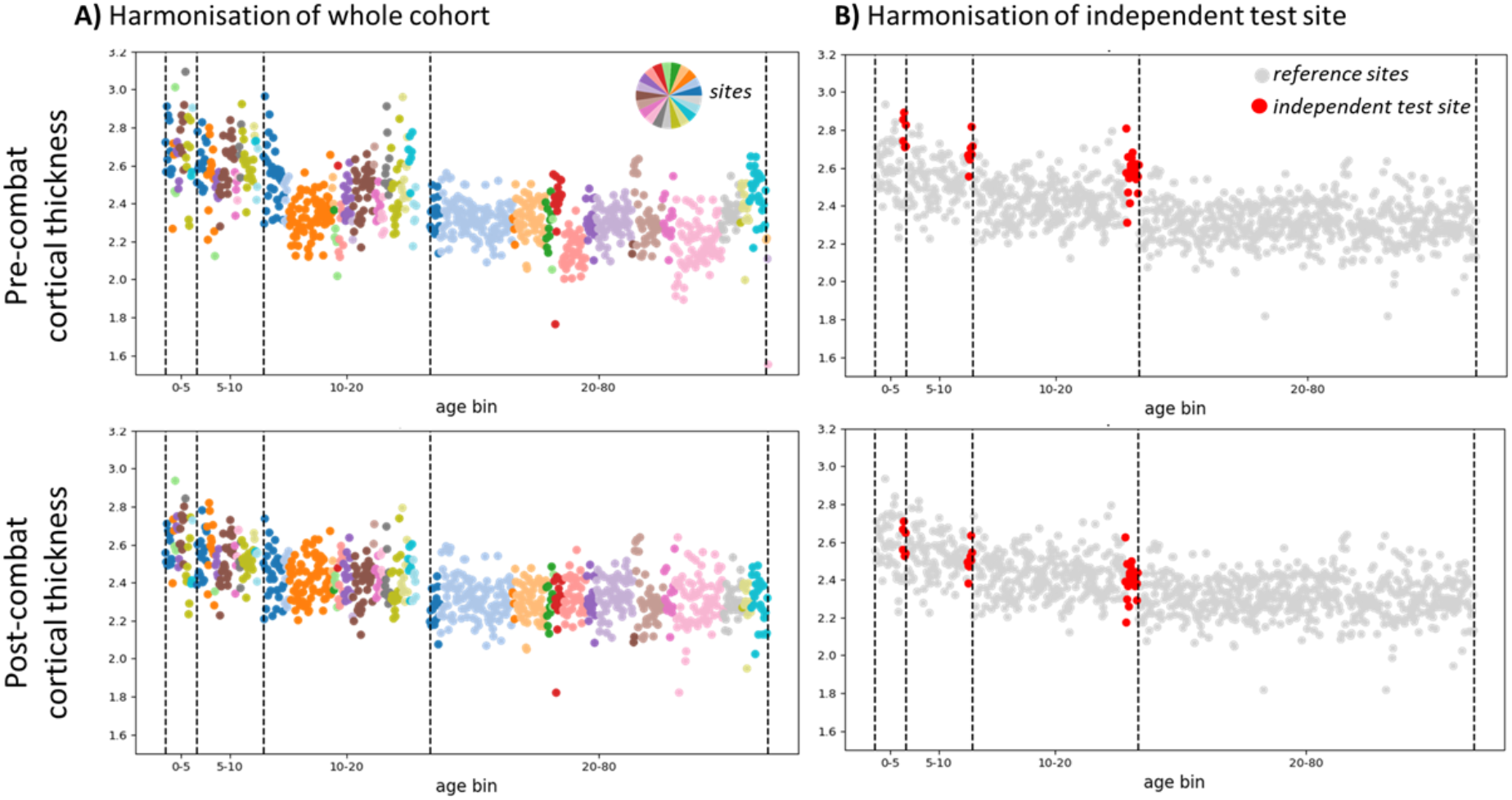
Multi-centre harmonisation of cortical thickness using ComBat. A) Pre- and post-combat cortical thickness of the whole cohort arranged per age bin and colored per site. Pre-combat site-differences in cortical thickness are evident. Post-combat site-differences are minimised, but biological variability (e.g. age) remains - cortical thickness decreases with age. B) Cortical thickness of an independent test site (red) was harmonised with the whole cohort’s post-combat cortical thickness (grey) as a reference. This enables new sites to use the classifier on their MRI data.

#### 2.5.2. Three stage normalisation of features

1. To account for interindividual differences, e.g. age-related changes, features were normalised using within-subject *z-*scoring. To account for intraindividual, regional variability in features, two further normalisation steps were carried out: interhemispheric asymmetry and intrasubject normalisation.
2. Inter-hemispheric asymmetry maps of features were created subtracting right hemisphere vertex values from left hemisphere values and vice versa.
3. The outputs from steps 1 and 2, were z-scored by distribution of features at each vertex from healthy controls to adjust for regional variability.

The output of these normalisation steps is a set of intrasubject and intersubject normalised features (henceforth ‘normalised’ features) and a set of intrasubject, asymmetry and intersubject normalised featured (henceforth ‘asymmetry’ features).

#### 2.5.3. Characterisation of FCD features on MRI

Surface-based morphological features were calculated within the lesion masks of all patients. For controls, data were sampled from similarly-sized regions for comparison. T1-derived features, available in all subjects, underwent UMAP embedding (McInnes et al., 2018), a non-linear dimensionality reduction where similar examples are plotted closer together. Lesions were clustered into groups according to their UMAP locations using a Gaussian mixture model.

#### 2.5.4. Border zones

Lesion masks were drawn conservatively, to maximise the proportion of lesional vertices within the mask. There is inherent uncertainty in the precise borders of manually delineated lesion masks. Feature abnormalities extended approximately 40mm beyond the lesion (Figure 3). To account for this uncertainty, border zones were created around each lesion mask extending 20 and 40mm across the cortical surface. Vertices between 0mm and 40mm from the lesion mask were excluded from training to reduce training on mislabelled data. Predicted lesion clusters within 20mm of the lesion masks classified as detected for the sensitivity+ metric (see network evaluation section).

**Figure 3.**
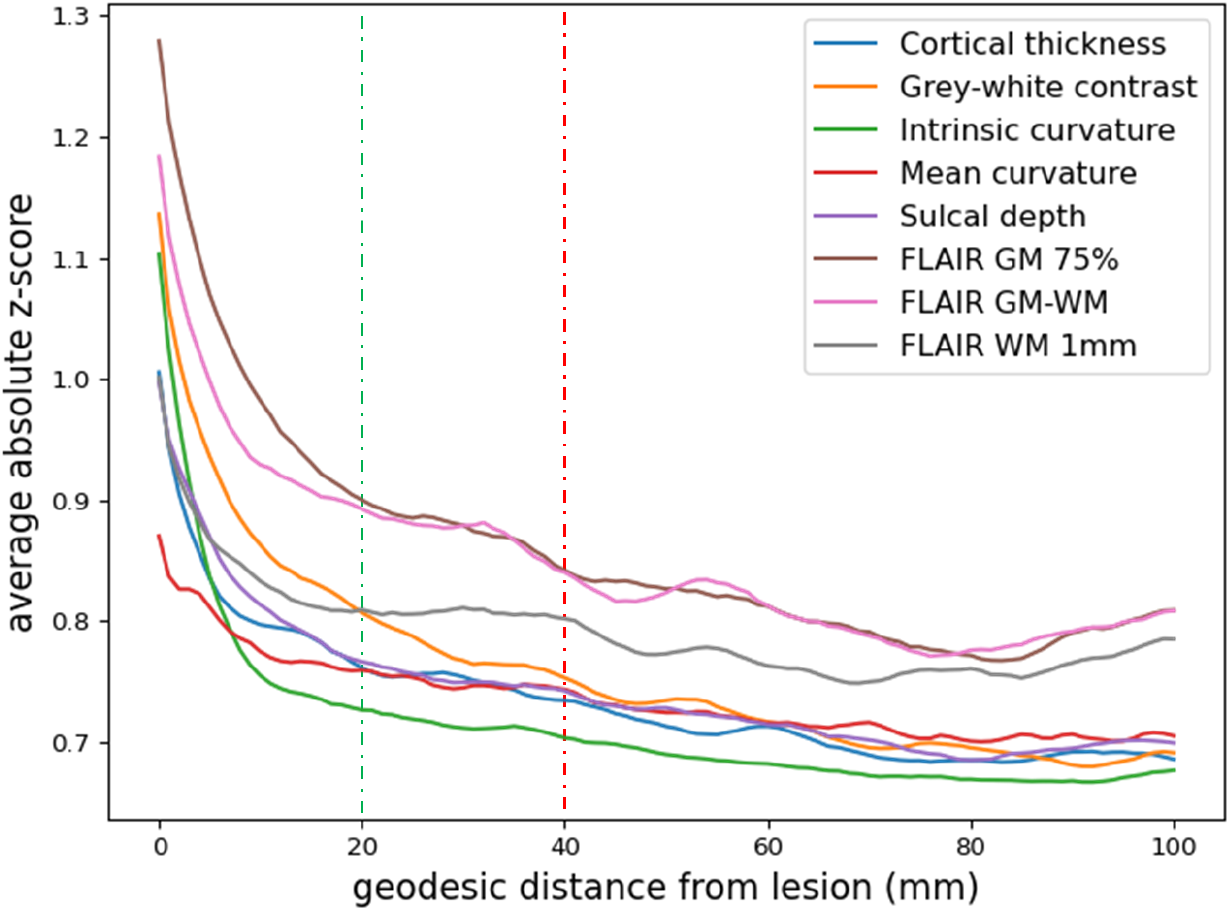
Extent of lesional abnormalities beyond the manual lesion masks. Normalised feature values plotted as a function of the geodesic distance from the manual lesion mask. Abnormal feature values extend up to 40mm (red dotted line) outside the manual lesion mask. From a distance of 40mm beyond the manual lesion mask, feature values look normal (red dotted line). To reduce data and label noise, during classifier training non-lesional samples are exclusively taken from beyond a 40mm border-zone, while lesional samples are all taken from within the manual lesion masks. For more precise estimates of classifier performance, any predicted lesion cluster within the 20mm border-zone (green dotted line) is also reported in the results.

### 2.6. Network Training, Testing and Interpretation

#### 2.6.1. Cohort splitting

An artificial neural network was trained on per-vertex post-processed MRI features [ComBat, Asymmetry, and Normalised], after border zones had been removed (33 total input features). The full cohort (excluding two independent test sites) of patients and controls were randomly assigned to either the train cohort (278 patients, 180 controls) or the test cohort (260 patients, 193 controls) (Table 1). All experiments to determine the optimal data processing and network parameters were carried out through 10-fold cross validation on the train cohort.

**Table 1.** Demographics table.

|  | Train cohort<br>Patients (n= 278) | Train cohort<br>Controls (n= 180) | Test cohort<br>Patients (n= 260) | Test cohort<br>Controls (n= 193) | Independent site 1<br>Patients (n= 17) | Independent site 1<br>Controls (n= 18) | Independent site 2<br>Patients (n= 16) |
| --- | --- | --- | --- | --- | --- | --- | --- |
| Age at preoperative scan<br>(median,IQR) | 20.0 , (11.0 - 32.8) | 29.0 , (19.0 - 37.9) | 18.0 , (11.0 - 29.0) | 29.0 , (19.5 - 39.2) | 7.3 , (5.2 - 11.1) | 14.6 , (10.5 - 16.1) | 6.1 , (3.4 - 16.2) |
| Sex (f:m) | 150 : 127 | 105 : 75 | 125 : 135 | 104 : 88 | 7 : 10 | 10 : 8 | 6 : 10 |
| Age of epilepsy onset<br>(median,IQR) | 6.0 , (2.5 - 12.0) |  | 6.0 , (3.0 - 11.0) |  | 3.4 , (0.8 - 5.8) |  | 2.0 , (0.9 - 5.1) |
| Duration of epilepsy<br>(median,IQR) | 10.0 , (4.3 - 18.4) |  | 10.2 , (5.0 - 18.2) |  | 3.0 , (0.5 - 7.2) |  | 2.4 , (1.3 - 8.1) |
| FLAIR available | 132 / 278 (47.0%) | 28 / 180 (16.0%) | 110 / 260 (42.0%) | 28 / 193 (15.0%) | 17 / 17 (100.0%) | 18 / 18 (100.0%) | 16 / 16 (100.0%) |
| Scanner (1.5T:3T) | 41 : 237 | 18 : 162 | 56 : 204 | 15 : 178 | 0 : 17 | 0 : 17 | 0 : 16 |
| Surgery | 208 / 278 (75.0%) |  | 190 / 260 (73.0%) |  | 5 / 17 (29.0%) |  | 16 / 16 (100.0%) |
| Histology | 193 / 208 (93.0%) |  | 171 / 190 (90.0%) |  | 4 / 5 (80.0%) |  | 16 / 16 (100.0%) |
| Seizure free | 123 / 183 (67.0%) |  | 106 / 157 (68.0%) |  | 3 / 5 (60.0%) |  | 14 / 16 (88.0%) |
| Follow up time | 2.0 , (1.0 - 3.0) |  | 2.0 , (1.0 - 3.4) |  | 1.5 , (1.1 - 1.7) |  | 2.9 , (1.9 - 4.4) |

**Table 2.** Classifier performance. Performance of the classifier on the test cohort, full cohort and the two independent sites.

|  | Sensitivity+<br>(percentage of<br>patients detected) | Sensitivity<br>(percentage of<br>patients detected) | Number of clusters in<br>patients<br>(median<br>(IQR)) | Specificity (percentage<br>of controls with zero<br>clusters) | Number of clusters in<br>controls (Median<br>(IQR)) |
| --- | --- | --- | --- | --- | --- |
| **Test cohort** | 67% (174/260) | 59% (154/260) | 2 (1.0-3.0) | 54% (105/193) | 0 (0.0-1.0) |
| **Full cohort** | 65% (350/538) | 58% (314/538) | 2 (1.0-3.0) | 52% (194/373) | 0 (0.0-1.0) |
| **Independent site 1** | 94% (16/17) | 88% (15/17) | 2 (2.0-4.0) | 17% (3/18) | 1 (1.0-2.0) |
| **Independent site 2** | 62% (10/16) | 56% (9/16) | 2 (2.0-3.25) | NA | NA |

#### 2.6.2. Network hyperparameters and training

The neural network architecture had 2 hidden layers (40, 10) and 1 output node and used a dropout of 0.4 on the input layer for learning more robust representations. To adjust for the class imbalance between healthy and lesional examples, for each patient 2000 random lesional and non-lesional vertices were sampled per epoch. If a patient had less than 2000 lesional vertices, existing lesional vertices were randomly drawn multiple times. A focal loss (Lin et al., 2020) was used to concentrate network training on difficult examples. After training, the network predictions were thresholded using an optimal threshold determined based on the Dice (F1) score on the train cohort. For the full list of optimised parameters see Supplementary Table 1.

On each of the 10 folds in the train cohort, a classifier was trained 5 times with different random initialisations. The resulting 50 models were combined into an ensemble model by averaging the individual models’ predictions, and evaluated on the test cohort. To calculate individual performance statistics for subjects in the train cohort, a second ensemble network was trained in a similar manner on the test cohort and evaluated on the train cohort.

#### 2.6.3. Evaluation metrics

Per-vertex lesion predictions for each individual were grouped into spatially-connected clusters on the surface-mesh. Clusters smaller than 100 vertices (approximately 0.5cm^2^) were filtered out as noise. The following outcome measures were calculated:

1. sensitivity, defined as the proportion of patients where a predicted lesion cluster overlapped the manual lesion mask;
2. sensitivity+, defined as the proportion of patients where a predicted lesion cluster overlapped the manual lesion mask or the border zone;
3. specificity, defined as the proportion of controls with zero clusters;
4. average number of clusters per patient;
5. average number of clusters per control.

#### 2.4.6. Network performance evaluation

Three complementary methods to understand and interrogate classifier performance and behaviour were used.

1. To determine how demographic and clinical factors influenced whether lesions were successfully detected by the classifier, two logistic regression models were used. The first included presurgically available variables: sex, scanner field strength, lesion hemisphere, FLAIR availability. The second included post-surgical variables (histopathological diagnosis and seizure freedom) and was applied on the cohort of patients who had undergone surgery. Statistical significance was determined through repeating regression analysis on randomly permuted cohorts (1000 permutations). Correction for multiple comparisons used the Benjamini-Hochberg procedure.
2. To understand classifier predictions, MRI features from predicted clusters were transformed into the UMAP embedded space described above.
3. To understand which specific features drove network predictions, integrated gradients saliency was computed (Sundararajan et al., 2017). This method computes which features are important to the network by looking at the integral (Riemann approximation) of the gradients computed from a baseline input (0 for each feature) to the actual feature values for each vertex.

### 2.7. Code and data availability

All data analysis was performed in Python. All protocols and code are available to download from <u>protocols.io/researchers/meld-project</u> and www.github.com/MELDProject/meld_classifier.

## 3. Results

### 3.1. Participant demographics

After excluding patients with missing lesion labels (n=37) and outliers (n=14), a total of 571 FCD patients were included (Table 1). Each epilepsy surgery centre contributed 6 to 87 patients. 419 patients underwent surgical intervention (73%) and histopathological diagnosis was available in 384 patients (92% of operated patients). Post-surgical outcome data was available in 361 patients (86% of operated patients); 68% were seizure free (Engel class 1) at last follow-up (median follow up = 2 years).

### 3.2. FCD lesion characterisation

UMAP embedding of surface-based features from manual lesion masks and equivalent healthy cortex in the full cohort is shown in Figure 4A. Compared to healthy control cortex, many lesions exhibited a distinct set of MRI features. There was heterogeneity in the set of abnormal features, with three distinct groups emerging (Figure 4B). Group 1 was predominantly composed of FCD type IIA, IIB and unoperated lesions. These lesions were generally located at the bottom of a sulcus and characterised by increased intrinsic curvature, increased cortical thickness, decreased grey-white matter contrast and increased FLAIR in the white matter. Group 2 lesions were characterised by increased intrinsic curvature, decreased grey-white matter contrast and decreased intracortical FLAIR. Group 3 lesions, in which the lesional features overlapped with healthy cortex, were more heterogeneous and had less extreme feature values.

**Figure 4.**
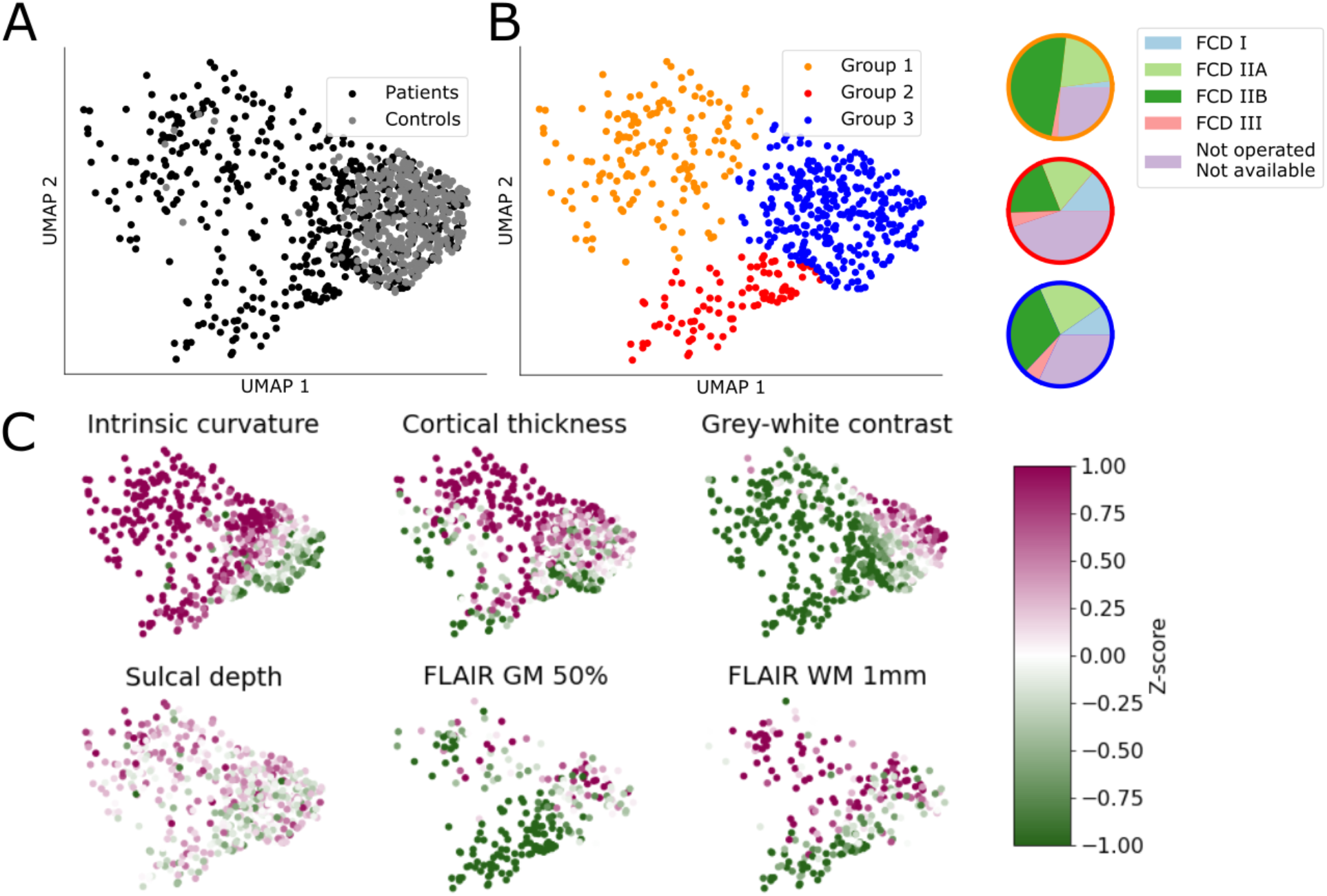
Non-linear 2D UMAP embedding of lesional T1 features. A) Manual lesion masks of patients (black), compared to equivalent cortex on healthy controls (grey). Lesions differ from control cortex and exhibit different patterns of structural abnormality. B) Data-driven clustering of UMAP embedding reveals 3 distinct groups of lesions. Colored-associated pie charts describe the proportion of each histopathological subtype present in each group. C) Patient lesions coloured by intra and inter-subject normalised features. Group 1 is predominantly FCD IIA & IIB, along with unoperated patients. It is characterised by increased intrinsic curvature, increased cortical thickness, decreased grey-white matter contrast, bottom of sulcus and increased FLAIR in the white matter. Group 2 is characterised by increased intrinsic curvature, decreased grey-white matter contrast and decreased intracortical FLAIR. It contains proportionally more FCD I and III lesions. Group 3 largely overlaps healthy control clusters. Lesional features in this cluster are more heterogeneous and less extreme.

### 3.3. Classifier performance

#### 3.3.1. Detection in the test cohort

For the 260 patients in the test cohort, the classifier predicted a median of 2 (interquartile range: 1 - 3) clusters. These clusters overlapped with the manual lesion mask in 154 patients (sensitivity = 59%), and overlapped with the extended lesion mask (including border-zones) in 174 patients (sensitivity+ = 67%). For the 193 controls in the test cohort, the classifier predicted a median of 0 (interquartile range: 0 - 1) clusters. No cluster was predicted in 105/193 controls (54% specificity). Examples of individual predictions for detected and undetected lesions are presented in Figure 5.

**Figure 5.**
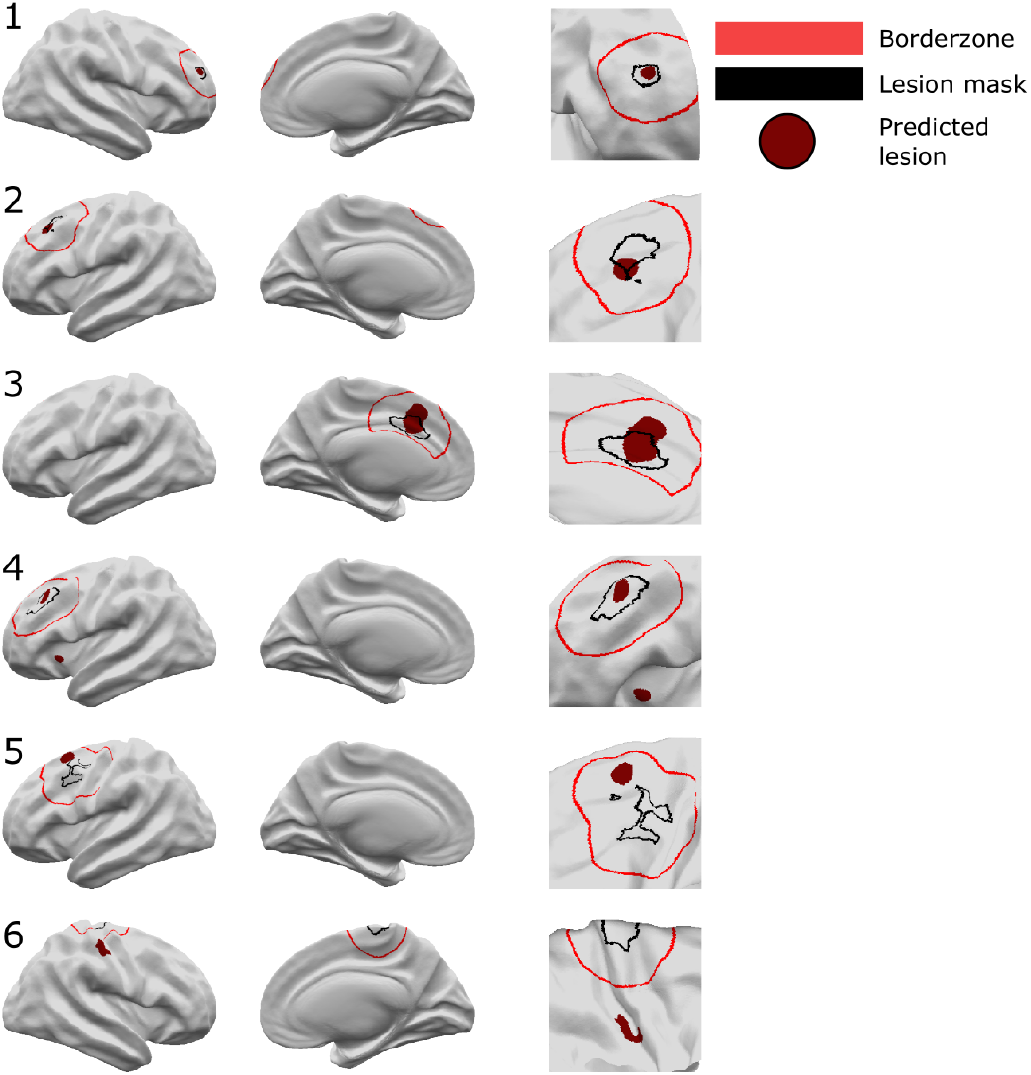
Neural network predictions. Classifier predictions for 6 patients are displayed. Patients 1-4 are examples where the classifier has correctly identified the lesion. In patient 4 there is an additional cluster in the left insula. Patient 5 is an example where the classifier detects an area in the border-zone. Patient 6 is an example of where the neural network has not identified the lesion. An additional cluster is detected in the right post central gyrus. Left column = lateral view, middle column = medial view, right column = zoomed in view around lesion mask. Black = lesion mask. Red = border-zone. Burgundy = classifier predicted clusters.

#### 3.3.2. Detection in the full cohort

In the full cohort (538 patients, 373 controls), i.e including predictions from training the network on the test dataset and testing on the train dataset, results were similar to those on the test cohort only. Sensitivity was 58%, sensitivity+ was 65% and specificity was 52%. The classifier predicted a median of 2 clusters in patients and 0 clusters in controls. Out of the 178 patients who were “ever reported MRI-negative”, clusters overlapped with the extended lesion mask (including border-zones) in 112 patients (sensitivity+ = 62.9%). On a restricted cohort of patients with T1 and FLAIR data, who had histopathologically confirmed FCD type IIB and were seizure free, sensitivity was 85%. Classifier performance according to histopathology is presented in Table 3.

**Table 3.** Classifier performance grouped according to demographic factors. Detection rate per age group, MRI status, seizure freedom, scanner strengths, MRI modality, and histopathology.

|  |  | % Detected | Patients (n) |
| --- | --- | --- | --- |
| **Age group** |  |  |  |
| Adult |  | 62.4 | 282 |
| Paediatric |  | 68.0 | 256 |
| **Ever reported MRI negative** |  |  |  |
| Visible |  | 66.1 | 360 |
| MRI negative |  | 62.9 | 178 |
| **Seizure freedom** |  |  |  |
| Seizure free |  | 69.9 | 229 |
| Not seizure free |  | 58.6 | 111 |
| **Scanner** | **Sequence** |  |  |
| 1.5T | T1 Only | 46.0 | 63 |
|  | T1 & FLAIR | 82.4 | 34 |
| 3T | T1 Only | 63.9 | 233 |
|  | T1 & FLAIR | 69.2 | 208 |
| **Histology** |  |  |  |
| FCD I |  | 50.0 | 44 |
| FCD IIA |  | 64.6 | 113 |
| FCD IIB |  | 76.8 | 185 |
| FCD III |  | 72.7 | 22 |
| Not available |  | 55.7 | 174 |
| **Restricted cohort** |  |  |  |
| T1 & FLAIR, FCD IIB, seizure free |  | 85.0 | 40 |

#### 3.3.3. Detection on independent test sites

When testing the classifier on the two independent sites, sensitivity was 88% for site 1 (sensitivity+ 94%) and 56% for site 2 (sensitivity+ 62%). Specificity for site 1 was 17%, lower than expected compared to the full cohort. However, the sample size of controls for this site was only 18.

### 3.4. Evaluating network performance across the full cohort

#### 3.4.1 Demographic and clinical factors affecting network sensitivity

The first logistic regression model (Figure 6A), based on pre-surgical factors, showed that lesions were more likely to be detected in patients who were operated (β=0.43, p=0.04) and those that had FLAIR data available if they were scanned on a 1.5T MRI scanner (β=1.10, p=0.01). Lesions were less likely to be detected in patients scanned on 1.5T scanners (β=-0.60, p=0.02) and when located in the left hemisphere (β=-0.41, p=0.02). However, these did not survive correction for the number of factors in the logistic regression model. There was no association with age, i.e. there was no significant difference in detection rates between paediatric and adult patients. Among post-surgical factors (Figure 6B), detection rates differed across histopathological subtypes, with 76.8% of FCD type IIB lesions detected, 64.6% of FCD type IIA, 72.7% in FCD type III and only 50.0% in FCD type I. FCD type I was significantly less likely to be detected (β=-0.53, p=0.01), and FCD 2B being more likely (β=0.57, p=0.02) than other histologies. Detection rates were non-significantly positively associated with post-surgical seizure freedom (β=0.51, p=0.04). Patients that are not seizure free may have more subtle lesions, which may contribute to both incomplete resections and the classifier not being able to detect them. Alternatively the lesions in patients that are not seizure free, may have been incorrectly masked.

**Figure 6.**
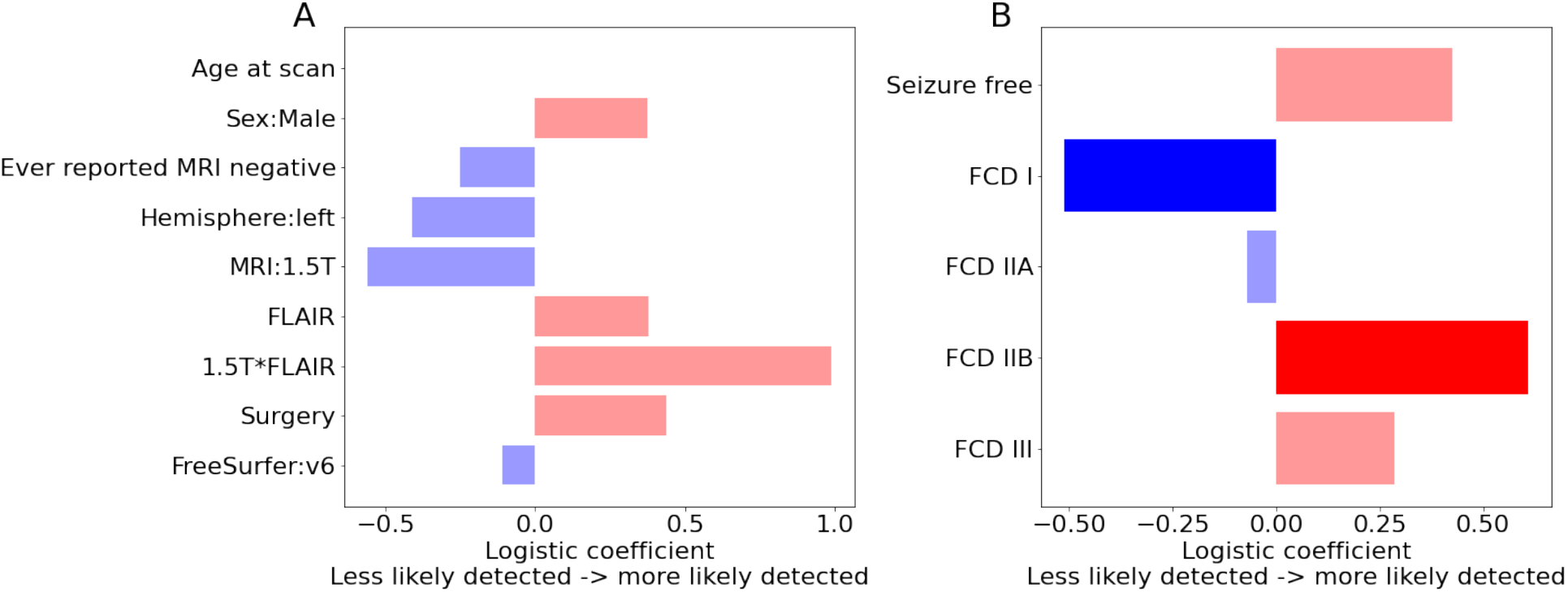
Logistic regression to determine factors associated with lesion detection. A) Presurgical factors and B) post-surgical factors associated with lesion detection. Bold colours indicate significance after correction for multiple comparisons. FCD IIB lesions were significantly more likely to be detected (bold red) and FCD I lesions were significantly less likely to be detected (bold blue).

#### 3.4.2. MRI features of predicted lesion clusters

The MRI features within the manually defined lesion masks clustered into 3 distinct groups (Figure 7A). Groups 1 and 2, were associated with high detection rates (96.0% and 82.8% respectively), whereas group 3, which largely overlapped healthy cortex, had much lower rates of detection (56.3%). A lower percentage of operated patients in group 3 were seizure free (59.0% compared to 78% in groups 1 and 2). Predicted lesion clusters superimposed on this UMAP embedding entirely overlapped groups 1 and 2 (Figure 7B) and no predicted lesion clusters were similar to group 3, which was indistinguishable from healthy cortex. For those manual lesion masks in group 3 that were correctly detected, the predicted lesion clusters exhibited features closer to those in groups 1 or 2 (Figure 7C). This indicates that while the manual lesion masks for lesions in group 3 did not capture areas of cortical surface that exhibited characteristically abnormal MRI features, the neural network learned to identify an overlapping set of vertices that did exhibit abnormal feature characteristics.

**Figure 7.**
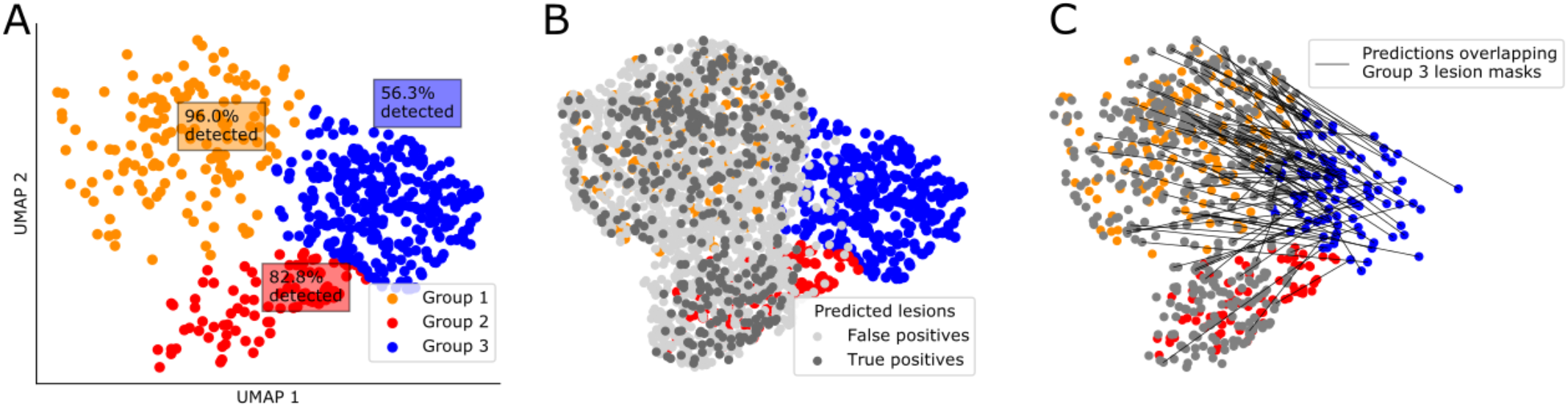
A) Data-driven clustering of UMAP embedding of lesional T1 features reveals 3 distinct groups of lesions. B) True positive and false positive clusters derived from the neural network superimposed on A. Feature values in true positive and false positive clusters are similar to either Group 1 or 2. Clusters are not similar to healthy cortex or group 3. C) Predicted clusters overlapping lesion masks from Group 3 lesions are superimposed. The feature values in the predicted clusters are similar to Group 1 or 2, i.e. the network has identified vertices exhibiting characteristically abnormal MRI features in FCD.

#### 3.4.3. Characterising features salient to the network in segmenting FCD lesions

In all patients, mean feature values and network saliencies were calculated for each feature within the predicted cluster. This enables the creation of a patient specific report containing the predicted lesion location, which features are abnormal within that predicted cluster and how much weight those features had in driving the classifier prediction, which we illustrate in Figure 8 with 2 examples. Patient 1’s predicted lesion has decreased FLAIR in the grey matter, blurring at the gray-white matter boundary on T1, and moderately increased intrinsic curvature (Figure 8B). From these features, the computed saliency scores indicate that the neural network considers the decreased grey matter FLAIR and grey-white contrast most important for its prediction of lesional vertices. Patient 2 is an example of an FCD type IIB lesion without FLAIR features (Figure 8). The predicted lesion has high intrinsic curvature, high cortical thickness and low grey-white matter boundary contrast. These are also the three features with positive saliency scores, i.e. feature values driving the classifier’s “lesion” prediction.

**Figure 8.**
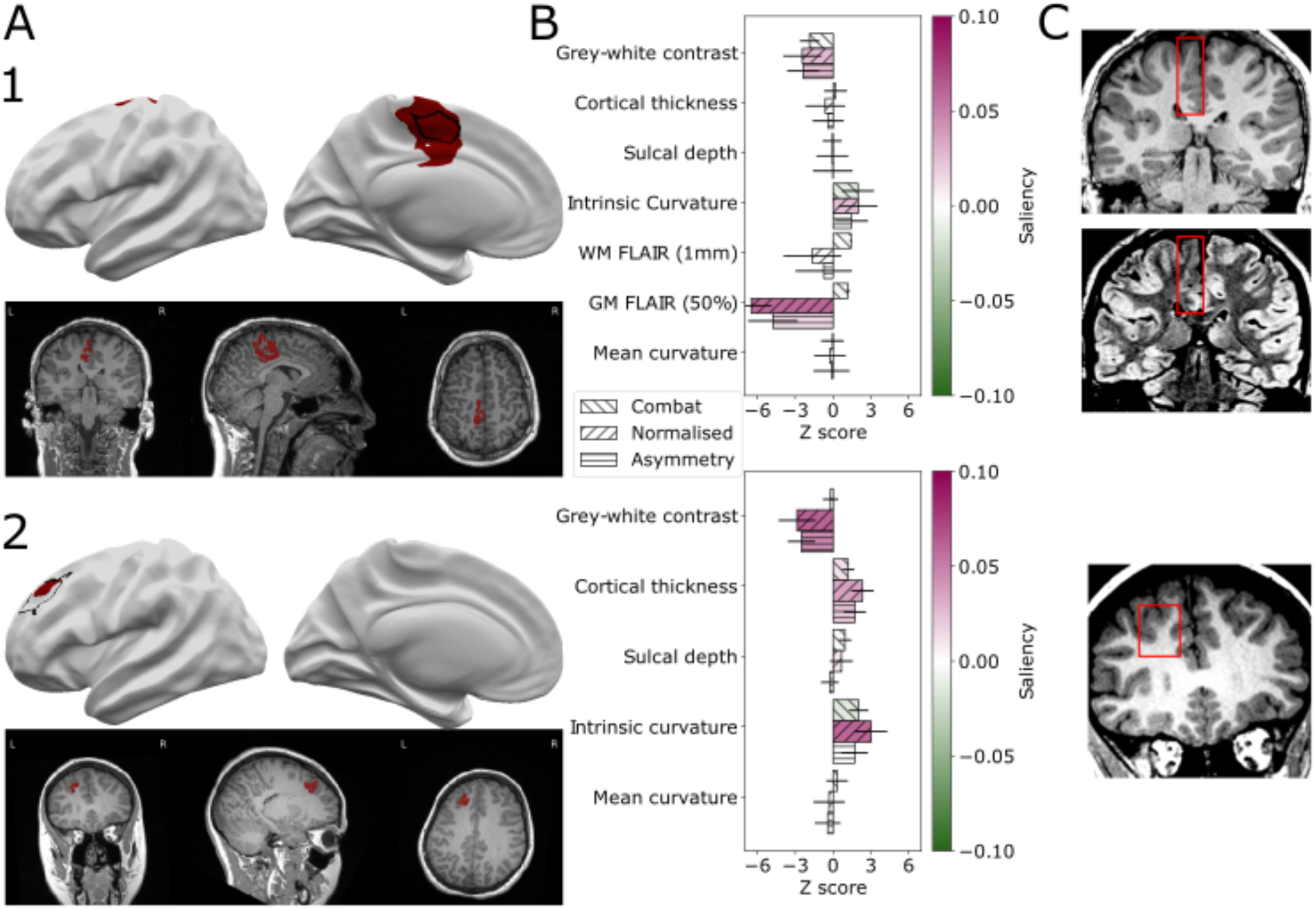
Individual patient reports. Example classifier predictions with saliency scores for “patient 1” (an example with FLAIR data) and “patient 2” (without FLAIR data). A) Classifier predictions (dark red) and manual lesion mask (black line) visualised on brain surfaces (only lesional hemisphere is shown). Classifier predictions (dark red) visualised on T1 volume. B) Z-scored mean feature values within predicted lesions colored with Integrated Gradients saliency scores. Positive saliency scores indicate feature values driving the classifier’s “lesion” prediction. Negative scores indicate feature values that are inconsistent with the prediction. C) Lesional cortex highlighted on the patients’ MRI scans exhibit salient features automatically identified by the classifier.

## 4. Discussion

We present an interpretable, fully automated pipeline for surface-based detection of focal cortical dysplasias, which has been validated on a large withheld test cohort, incorporating data from 20 sites, and two independent sites. The sensitivity to detect lesions in the test cohort was 67%, with sensitivities of 94% and 62% in the independent sites, 85% in the restricted cohort and 63% within patients who were ever reported “MRI-negative”. Logistic regression analyses indicated that FCD type IIB lesions had higher detection rates, whereas FCD type I lesions had lower detection rates. Multidimensional analysis of lesional cortex revealed groups of lesions characterised by different MRI features, histologies, post-surgical outcomes and detection rates. Individual patient reports provide a map of the predicted lesion locations alongside the quantified lesional features and how salient they were considered by the classifier.

This study extends previous work on FCD detection in the largest MRI cohort of FCDs to date. Previous surface-based work has identified features that differentiate lesional cortex and developed machine-learning frameworks for the incorporation of these features (Adler et al., 2017; Hong et al., 2014; Jin et al., 2018; Mo et al., 2018; Wagstyl et al., 2020). However, being limited by small numbers of patients and data acquired from only one or two MRI scanners can lead to large error bars on estimates of sensitivity and specificity (Varoquaux, 2018) and limited generalisability due lack of diversity in training data. Progress is also being made on automated volumetric MRI methods (David et al., 2021; Gill et al., 2021; House et al., 2021), however datasets have tended to be restricted either to histopathologically confirmed FCD type II cohorts, or single sites. Through creating a large dataset including both paediatric and adult patients across multiple sites and MRI scanners as well as including all FCD histopathological subtypes, we aimed to address these limitations. Furthermore, we included lesions that were visible to neuroradiologists on MRI as well as those that had been reported “MRI-negative”. Lastly, our training dataset included lesions masked by different radiologists / researchers at different institutes. This heterogeneity in lesion masking reduced overfitting of the network to one individual neuroradiologist’s opinion. This large multisite, multiscanner cohort provided reliable and reproducible estimates of classifier performance that generalised well to two independent cohorts. In particular, it was able to detect 76.8% of FCD type IIB lesions.

Our data-driven clustering of FCD lesions revealed three distinct groups of lesions. Group 1 had “classical” radiological features of FCD type II; increased cortical thickness, blurring of the grey-white matter boundary, abnormal folding, FLAIR hyperintensity in the white matter and were often located at the bottom of sulci. They were associated with high detection rates by the neural network (96%) and had good seizure freedom rates (78%). Group 2 had more subtle features; blurring of the grey-white matter boundary, FLAIR hypointensity in the grey matter and some folding changes. However, our classifier was still able to detect 82.8% of these lesions and the patients in this group who had been operated on still had good seizure freedom rates (78%). In contrast, lesions in group 3 were difficult to differentiate from healthy cortex, they did not demonstrate characteristic FCD “fingerprints” and only 59% of these patients were seizure free after surgery. For group 3 lesions that were detected by the classifier (56.3%), the classifier identified a subset of vertices that exhibited MRI features more consistent with groups 1 and 2 (Figure 4C). This suggests that these lesions are more subtle or difficult to delineate or structurally heterogeneous (Lee et al., 2020) on MRI.

One challenge in incorporating machine-learning algorithms in clinical practice is their perception as being “black-boxes”, with limited feedback on what data have informed a prediction. Saliency aims to interrogate which specific input features drive neural network predictions. Our individual patient reports provide information on which features are abnormal within the predicted clusters, accompanied by their impact on classifier prediction (Figure 8). A neuroradiologist or multidisciplinary team could use this tool to confirm their hypotheses in “MRI-visible” lesions, to rereview the scans of “MRI-negative” patients or identify targets for more detailed investigations, such as sEEG (Wagstyl et al., 2020). They will obtain putative lesion locations identified by the classifier, equipped with an understanding of what features were considered suspicious and how they were abnormal, thus opening the “black-box”.

## 5. Limitations and future work

Participating MELD sites manually masked FCD lesions and only surface-based data were shared with the project coordinators. While preserving a greater level of anonymity and facilitating data sharing, this preprocessing prevented assessment of inter-rater reliability of lesion masks and comparison of predicted lesions with patients’ volumetric MRIs. As with other FCD detection algorithms, false positives were common in both patients and controls. This neural network classifies individual vertices, future work using incorporating neighbourhood information and incorporation with volumetric approaches may help to reduce the false positives. Additionally, integrating electrophysiology might help to identify which structural abnormalities are epileptogenic. One challenge in all FCD detection work is deciding which patients are considered “MRI-negative”. The measure “ever reported MRI-negative” will vary based on the level of neuroradiological expertise at the individual site as well as the MRI scanner and sequences acquired. However, it should provide a measure of the more challenging lesions to detect. Lastly, drug-resistant focal epilepsy is caused by multiple pathologies of which FCDs are a significant subset. Invaluable future studies would extend the inclusion criteria to a wider spectrum of focal epilepsies.

## 6. Conclusions

We demonstrate how through open-science practices and de-centralised MRI post-processing, one can create a dataset; and train and validate a machine-learning framework to assist in the diagnosis of a rare clinically-challenging pathology. The MELD FCD classifier is a fully automated, open-access surface-based tool that can be run on any patient with a suspicion of having an FCD who is over age three and has a 1.5T or 3T T1 scan, with or without FLAIR data. The classifier is available on GitHub as a user-friendly python package and can output a patient specific report detailing suspected structural abnormalities, which features are abnormal within these clusters and their impact on classifier prediction.

## Data Availability

Code to reproduce all analyses and figures in this manuscript is available to download from github.com/MELDproject. Requests can be made for access to the full MELD dataset.

https://www.protocols.io/researchers/meld-project

## 9. Funding and acknowledgements

The MELD project is supported by the Rosetrees Trust (A2665). We are grateful to ENIGMA-Epilepsy for paving the way for collaborative neuroimaging cohorts in epilepsy and open protocols. This work is supported by the NIHR GOSH BRC. The views expressed are those of the authors and not necessarily those of the NHS, the NIHR or the Department of Health. KSW is supported by the Wellcome Trust (215901/Z/19/Z). NTC is supported by the CNF/PERF Shields Award, and the CNRI Chief Research Officer Award. XY, NTC & WDG are supported by the Hess Foundation and Children’s National IDDRC. FC and CY were supported by the São Paulo Research Foundation (FAPESP), Grant # 2013/07559-3 (BRAINN - Brazilian Institute of Neuroscience and Neurotechnology). JOM is supported by a Sir Henry Dale Fellowship jointly funded by the Wellcome Trust and the Royal Society (Grant Number 206675/Z/17/Z) and received support from the Medical Research Council Centre for Neurodevelopmental Disorders, King’s College London (grant MR/N026063/1). JJM and KZ are funded by the National Natural Science Foundation of China (No. 82071457). PS acknowledges the DINOGMI Department of Excellence of MIUR 2018-2022 (legge 232 del 2016). GPW is supported by the MRC (G0802012, MR/MR00841X/1). KJW is supported by The Alan Turing Institute under the EPSRC grant EP/N510129/1. IW is supported by NIH R01 NS109439. RG and CB are supported by Tuscany Region Call for Health 2018 (grant DECODE-EE);. JD is supported by NIHR and Wellcome Trust (218380). TJO, LV, AW and BS were supported by an NHMRC Investigator Grant #APP1176426. AC is supported by a GOSH Children’s Charity Surgeon-Scientist Fellowship. RK and YL have been funded by the Saastamoinen Foundation. KH and SD were supported as part of the BRAIN Unit Infrastructure Award (Grant no: UA05). The BRAIN Unit is funded by the Welsh Government through Health and Care Research Wales.

## Supplementary Materials

**Supplementary table.** List of neural network parameters.

| ***Input data selection*** |  |  |
| --- | --- | --- |
| Number of vertices per subject | 2000 lesional, 2000 non-lesional | If there are less than 2000 lesional vertices, they are oversampled. Non-lesional vertices are drawn randomly from a pool of 10000 vertices each epoch |
| Size of boundary zone | 40mm | Vertices from the boundary zone are excluded from training |
| Shuffle each epoch | True |  |
| Normalisation | Per-feature mean and std |  |
| ***Neural network architecture*** |  |  |
| Layers | Input (xx) - Dense (40) - Dense (10) - Output (2) |  |
| Dropout | 0.4 | Applied after each layer |
| ***Training*** |  |  |
| Loss | Focal loss, alpha=0.2, gamma=5 |  |
| Learning rate | 0.00001 |  |
| Batch size | 1024 |  |
| Epochs | 100 |  |
| Patience | 10 | Training was stopped early, if loss did not improve after 10 Epochs |
| Number of folds | 10 | Networks trained with 10-fold cross validation were ensembled to a final model |
| ***Evaluation*** |  |  |
| Optimal threshold | 0.52 | Computed on training set: |
| Min area lesion | 100 vertices | Minimum number of connected vertices predicted as lesion to be considered a predicted lesional cluster |

